# Development of a parent education module for screen-dependent preschool children with social communication deficit

**DOI:** 10.1101/2025.08.03.25332562

**Authors:** Joe Mathew V J, Kishan M M, Shivani Tiwari

## Abstract

Technology and its advancements have significantly impacted the quality of life for humans. With the progress in technology, screen dependency has also progressed in people of all ages. Despite available guidelines recent surveys indicate that the screen time in children is beyond the recommended levels. Excess screen time is known to create behavioural issues, delayed language development, poor expressive language, poor social skills, and traits like autism (aka virtual autism) in young children. Children with such traits demonstrate significant improvements in their social and language skills when screen time is reduced with adequate guidance and stimulation. The present study, thus, aimed to develop a parental education module to reduce screen time for preschool children with social communication deficits. At first, the contents of the module were developed through a parental in-depth interview conducted to find the issues faced by parents due to screen time in children, in addition to a focused literature search. A panel of experts rated the contents of the module in terms of relevance, age appropriateness and feasibility of implementation. Expert rating showed a high consensus (80%) among the raters and appropriateness of its contents. Furthermore, feedback received on the revised module revealed acceptance by the parents. The module is expected to improve the parental awareness, knowledge and attitude about the challenges of screen time in children and thereby control the effects of screen time in this group.

## Introduction

In the current technologically advanced era, newer developments in digital technology multi-fold the need for digital media usage. We use digital media in the form of mobile phones, tablets, iPads, computers, television, etc., and the purpose ranges from education, recreation, social communication, food, groceries, etc. Consequently, screen time and digital media usage are rising alarmingly in individuals of all ages [1]. ‘Screen time’, therefore, refers to the time spent by an individual in front of an electronic device having a screen like a television, tablet or a mobile phone [2]. COVID-19, caused by the coronavirus, which started in 2019, has spread worldwide and has had drastic effects worldwide. This resulted in tremendous changes in the lifestyle of the human population [3]. Isolation to restrict the spread of infection led people to depend greatly on digital media for communication, education, recreation, etc [4]. This caused a further increase in the screen exposure for individuals across age groups. A study that assessed the screen time changes in preschool children in China during the period of COVID-19, reported an increase of one hour for TV and video watching and half an hour for touch screen devices in preschool children during COVID-19 [5]. As per the authors, most of the screen time was utilized for the purpose of education and entertainment.

A recent meta-analysis compared the screen time in children and youth over the age range of 0-21 years before and during the COVID-19 pandemic across the globe [6]. This meta-analysis of 51 studies revealed that children and youth between 0-21 years spent an average of 2.67 hours (SD 0.21) per day on screen before the pandemic. The average screen time increased to 4.38 hours (SD 0.21) per day during the pandemic, based on the meta-analysis of 91 post COVID-19 disease studies. Children aged between 0 and 5 years had an average screen time of 1.91 hours per day. This is more than the maximum prescribed screen time in children of this age. During the pandemic, this number further increased to 2.65 hours per day. Hence, there was a rise in screen time by 1.35 hours per day. Boys were found to have longer screen time before the pandemic, but there was no significant change in the duration of screen time across genders during COVID-19 [6].

According to a recent survey conducted in India [7], among 50 parents of children below 4 years of age, electronic devices are used by 92% of them. About 72% of these children use electronic gadgets regularly for more than two hours daily, and 46% do so without parental supervision. While 80% of the parents encourage their kids to participate in outdoor activities, 78% of the children react negatively when electronic gadgets are restricted. Approximately 80% of the parents knew they should spend quality time talking to their children. However, only 18% of the parents admitted that their children are addicted to electronic devices. Only 12% of parents were aware of the term ‘virtual autism,’ while 88% of them believed that virtual autism is a form of autism [7].

Multiple factors could explain excess screen time in young children. Firstly, the parental factors include their belief that TV is an educational resource for children, which improves knowledge, and that such devices will help develop the vocabulary and language skills in children [8]. Owing to the parents’ busy work schedule, it helps babysit and calm down children. Screen devices also help in providing some rest time for parents themselves. There is no formal rule for allowed screen time at home. Furthermore, parents are not being good models for the screen. Parent’s device use at home further encourages children to explore and view these devices. Elder siblings and friends are also sometimes bad models for younger children at home regarding screen use [9].

Recent advancements in technology are increasingly making it necessary for everybody to be dependent on devices and their numerous applications in our daily lives. Advancements in the education system promote technology to teach children at school better. Children are compelled to use the screen to attend their classes online. They also use the internet to access references and resources for their education instead of going to the library or their teachers. It also helps students complete their assignments and find new ideas[10].

The increase in screen time among children is reported to have adverse effects on their health and well-being. Some of the ill effects of excess screen time include obesity, sleep disturbance, postural effects, visual disturbance, and impaired cognitive development [11]. According to a recent study [12], children exposed to excessive screen time, were reported to experience a decline in cognitive ability, impaired language development, and mood disorders. Additionally, such children may exhibit behaviours similar to those of children with autism, such as hyperactivity, short attention span, and irritability.

Excess screen time in children affects their language ability. Screen time without parental assistance deprives young children of naturalistic human interaction. This might reduce the need for communication in them, which in turn could result in speech and language delays [13,14]. A recent systematic review examined the relationship between screen time and language acquisition in children. The review reported that early screen exposure causes delayed language acquisition in children. Screen time without parental assistance was found to have 8.47 times greater effect on language acquisition, by delaying it when compared to screen time in children with parental assistance. Mealtime screen exposure is reported to result in delays in expressive language and verbal interaction patterns in children between 2 and 5 years of age [15]. Screen time also affects verbal expression and phonological processing skills [16]. An evaluation of the impact of screen exposure among toddlers in India demonstrated that children under two years of age who viewed screen for less than 3 hours a day, were found to have delayed language skills and short attention spans while those children who had screen time of more than 3 hours a day were found to have hyperactivity along with language delays and short attention spans. Furthermore, children under 12 months of age with screen time exposure greater than two hours per day were six times more vulnerable to experience speech delay [12].

Autism Spectrum Disorder (ASD) or autism, according to DSM-5, is a deficit in social communication or interaction affecting social-emotional reciprocity, nonverbal communication behaviour used for social interaction, and the deficit in developing, maintaining, and understanding relationships, along with other restricted and repetitive behaviours [17]. Children with ASD display restricted and repetitive patterns in behaviour, interests and activities. Research suggests that children with screen exposure before three years of age are vulnerable to demonstrate autism-like features, which include poor attention, delayed cognitive skills, hyperactive behaviour, and poor social skills [18]. Consequently, ‘virtual autism’ is a recent concept introduced for children below three years of age with an average of four hours of screen exposure per day (exceeding the recommended safe limit of screen exposure), which results in later behavioural and language issues that resemble autism [18]. Children with such traits were observed to show good improvement when the screen time was controlled, and they were engaged in meaningful social activities, while such an improvement was not observed in children with ASD [19]. A case report by Numata-Uematsu et al.[20] was the first official documentation in the medical literature describing a five-year-old boy without any significant developmental history but having had high screen exposure from six months of age. The child had poor eye contact and other neurobehavioral problems. The Childhood Autism Rating Scale (CARS) score was 37, indicating severe autism. On clinical observation, the child liked to play with others and was not sensitive to physical contact or sensory stimuli, which was dissimilar to autistic behaviour. The child improved in communication and social behaviours after two weeks of no exposure to media without any intervention. The child’s CARS score on evaluation after two months was reduced to 26.5, which indicated no autism.

Given the various detrimental effects of excessive screen time on children’s development, the World Health Organisation [21] has provided guidelines on the time allocation of physical activity, sedentary behaviour, and sleep in children below 5 years of age. It is recommended that children should not have any sedentary screen time up to 1 year of age. Children aged 2-4 years are advised not to have more than one hour of screen exposure. Parents are advised to engage children below one year of age in physical activity for at least 30 minutes (through floor-based activity), and a minimum of 180 minutes (including intense physical activity) in children between 1 and 4 years of age. It is further advocated to follow good quality sleep of 14-17 hours in infants up to 3 months, 12-16 hours in children between 4-11 months, 11-14 hours in children between 1-2 years, and 10-13 hours in children between 3-4 years of age.

American Academy of Paediatrics, Council on Communications and Media [22] recommends that children under two years of age should have hands-on exploration and social interaction with trusted caregivers to develop adequate cognitive, language, motor, and social-emotional skills. It is now known that increased screen time impacts body mass index, leading to obesity. Early childhood screen time exposure during bedtime or sleep affects the sleep duration and cycle. It also affects children’s cognitive, language, and social/emotional skills [23]. It is, therefore, recommended that children below 18 months should not be exposed to screens other than a video chat. While introducing screen time to children between 18 and 24 months, it is recommended that they view high-quality content with parental interaction. Thus, it is not prescribed to let children have screen time alone. Children above the age of 2 years should not have a screen time of more than 1 hour per day. It is further suggested that children should not have screen time during meals and at least one hour before sleep [23].

On similar lines, the Indian Academy of Paediatrics Guidelines [11] put forth that children up to 24 months of age should not be exposed to any type of screen. No screen time is recommended during feeding or to calm the child’s crying. The caretaker should avoid screen time while engaging with the child. Children between 24 and 59 months of age should have a maximum of 1 hour of screen time per day, with each session not exceeding 20 – 30 minutes. Parents should monitor children’s screentime. Furthermore, content watched by the children should be educational, age-appropriate, non-violent, healthy, and preferably interactive. Children between 5-10 years of age can have a maximum screen time of 2 hours with minimal recreational screen time and more educational, learning and social interaction [11].

Interestingly, some of the studies document positive effects of screen time when utilised appropriately. The literature suggests that children between 2 and 5 years can be provided minimal screen time with parental assistance. According to the position statement by the Canadian Paediatric Society, effective utilisation of screen time in children can create a positive impact on children [24]. For instance, parents should monitor their children’s screen time, i.e., ensure their children are viewing quality content. During screen viewing, there should be one-to-one conversation between the child and co-viewer(s). The effective use of screens can help children improve their vocabulary, communication, and literacy skills. It also improves cognitive skills and imaginative play. Well-designed, age-appropriate educational programs and screen activities can be powerfully pro-social, helping children to learn anti-violence attitudes, empathy, tolerance and respect [25, 26]. Screen time can also be helpful for the child’s attention, especially in medical procedures. Activity-based programs and computer applications can enhance physical activity in children. Studies show that video games did positively improve physical activity in 3- to 17-year-old children. It also improved their cognitive skill [27].

Technology and screens are part of our life as well as the future, and all domains of life (including education) are increasingly dependent on their use. It is therefore important to ensure the safe use of screens among children. One important step towards this is to work on parental awareness and to educate the parents on the effects of excess screen time and its impact on various domains of development in young preschool children.

Khoo et al. performed pre- and post-intervention assessments on parents of children diagnosed with ASD, who had a screen time exposure of less than 120 minutes and more than 120 minutes every day. Thirty children with an average age of 7.5 years were selected to identify the effectiveness of the parent training program and their social functioning skills. Twenty-eight parents participated in the follow-up session. Parent training was conducted based on the AAP recommendations on screen time for children and its effects. Pre-post assessment was done using the Social Responsiveness Scale, 2nd edition – SRS 2 [28]. Results showed a reduction in the screen time post-intervention in more than half of the children whose parents attended the training program. The participant who had a significant decrease in screen time post-intervention, the SRS score significantly improved, wherein the social communication sub-score improved significantly in children with reduced screen time post-intervention. Another parent intervention program was conducted on nine children aged between 18 and 40 months with a diagnosis of ASD and having a screen exposure of more than 2 hours per day [29]. Parent intervention was delivered for six months, which consisted of a baseline assessment of parents followed by a 40-minute informational video summarizing the association between screen time and developmental problems. The weekly support system used strategies to reduce screen time and improve social interaction. Child autism symptoms (Brief Observation of Social Communication Change; Grzadzinski et al., 2016) [30], functional behaviour (Vineland Adaptive Behavior Scales; Sparrow et al., 2016) [31], and developmental (Mullen Scales of Early Learning; Mullen, 1995) [32] assessments were carried out pre- and post-intervention to compare the differences. Parental stress (Autism Parenting Stress Index; Silva et al., 2012) [33] was also assessed both at pre- and post-intervention times. The results showed a significant decrease in the screen time from an average of 5.6 hours per day before intervention to 4.72 minutes per day during the six-month period of the study. Further, there was a significant improvement observed in core autism symptoms in the children and in parental stress from pre- to post-intervention [29].

Another study [34] attempted to find the effect of parent training on children between the ages of 2 and 4 years. The duration of screen time, repetitive behaviour (Repetitive behaviour scale-Revised, RBS-R) and brain electrophysiological characteristics (using EEG recording) were assessed pre- and post-intervention. Twelve children participated in the study, who had screen time for more than half of their waking hours, along with sub-threshold autistic features. Along with these measures, a lifestyle checklist was also administered pre- and post-intervention, and a follow-up assessment 2 months after the last intervention session. Behavioural results showed a significant reduction in screen time and reduced stereotypic behaviours in children with ASD, along with increased parent-child interaction. Electrophysiological results showed changes in values post-intervention, indicating the intervention has positively impacted the behaviour and created observable changes in brain activation patterns.

Kaur et al. [35] developed a multicomponent intervention program, called Program to Lower Unwanted Media Screens (PLUMS), for typically developing children and their parents. This program consisted of an intervention for parents based on self-determination theory and children based on social cognitive theory. The research was carried out in two phases. Phase 1 was the preintervention phase that focused on the actual development of PLUMS, which was carried out in three stages. First, an initial draft was developed based on the existing literature; second, the children’s primary caregivers and professionals from various disciplines suggested possible alternatives to reduce screen time. An expert panel refined the ideas gathered, and third, the PLUMS draft was finalised as two separate intervention modules for the parent and the child. In the third stage, the feasibility, acceptability, and adherence to the PLUMS intervention plan were pretested in ten families. The second phase of the study [36] checked for the administration of the PLUMS program on families within zone three of Chandigarh. The target group for this intervention was parents of children between 2 and 5 years of age. Participants were divided into two groups-intervention and control. Each group had 170 participants. The study population included families with children between 2 and 5 years of age. The module was delivered online as videos to parents and implemented for 8 weeks. There was an introductory video and seven theme-based videos, which were shared with parents each week. A module for the child was also shared with activities. A digital screen exposure questionnaire, preschool child behaviour checklist and sleep disturbance scale for children were administered during baseline, post-intervention and follow-up sessions. Results of the study indicated that there was a significant reduction in screentime post-intervention and follow-up after 6 months. Comparing the control and intervention groups, there was a significant reduction in screen time post-intervention in the intervention group, but no significant difference was observed during follow-up. The duration of physical activity significantly increased, with a significant difference in the duration of physical activity between the intervention and the control group during post-intervention and follow-up after 6 months. The child’s sleep and emotional behaviour did not show any significant changes with the intervention of PLUMS in both groups.

Another study [37] conducted in northeast Thailand examined the efficacy of a newly developed training program for parents to reduce screen time in typically developing preschool children. This study recruited parents of 67 preschool children between 2 and 5 years of age, with a screen time of over one hour per day. The experimental group, with 35 participants, received a two-week intervention program consisting of 3 sessions of three hours each. The control group, with 32 participants, received a child’s screen time handbook after completing data collection. The intervention program, along with six questionnaires developed by the primary investigator, was validated by experts, involving a paediatrician, a paediatric nursing instructor and an early childhood education instructor. The questionnaire was administered one week and two months after the completion of the intervention. The study findings indicate that parent’s attitudes and behaviour towards children’s screen time in the experimental group were significantly higher than those in the control group immediately after the intervention. Additionally, the average score of parents’ perceived behaviour control on child’s screen time reduction was significantly higher in the experimental group than in the control group, two months after the intervention.

The above-presented review of literature suggests that there is a significant reduction observed in the screen time for children diagnosed with and without ASD following an effective parental education program. Reducing screen time in children with ASD brings positive changes in social communication skills, parent-child interaction, reduction in stereotypic behaviour, and autism features, increased sensitivity to novelty and reduced parental stress. While in typically developing children, parent training improved the attitude, behaviour, and parents’ perceived behaviour control on the child’s screen time. It also reduced children’s screen time and improved the duration of physical activity.

Given the range of negative effects of excessive screen time on various developmental skills, including social and communication skills, there is an immediate need for parental educational programs to combat excessive screen time in young children who present with social communication deficits, similar to children with autism. The present study thus aimed to develop an educational module for parents of screen-dependent, typically developing preschool children who show social communication deficits.

### Methodology

The present study followed a combination of qualitative and quantitative research design. The study protocol was reviewed and approved by the Institutional Research Committee (IRC), followed by the Institutional Ethics Committee. The study was carried out between August 2024 to April 2025. The parental educational module development was carried out in the following steps:

### Content development

The contents for the parent educational module were identified by a literature search and a semi-structured interview conducted on six parents of preschool children aged 2 to 5 years, who had a screen exposure greater than one hour per day. Parents who spoke and understood English with ease were recruited for the interview. Participants were briefed about the study purpose, and written consent were taken before the interview. The interview aimed to identify the parental concerns regarding their screen-dependent children, for inclusion in the module. A few open-ended questions were prepared for the interview (provided in Appendix A). The questions in the interview addressed children’s screen time, awareness of parents regarding the effects of increased screen time, challenges faced in enforcing screen control, social communication and other behavioural difficulties faced by the child, if any. The semi-structured interview probed into various difficulties and challenges in the family due to excessive screen time. The responses from the parents were audio recorded. The responses were carefully heard offline and, transcribed verbatim, to generate codes, analyse and identify relevant themes.

### Item generation and Content validation

Based on the responses from the thematic analysis, focused review of literature and expert discussion, an initial draft of the educational module incorporating each of the needs with relevant home-based activities/routines/suggestions was prepared. These home-based activities were carefully selected by referring to the available guidelines [21, 38] and other relevant literature [39–41]. The draft module was then subjected to content validation by a panel of experts. The experts for content validation involved five health care professionals (paediatrician, psychiatrist, speech-language pathologist, occupational therapist, clinical psychologist) who dealt with developmental impairments, and a parent having a child aged 2-5 years with screen time above the recommended level (i.e., exceeding 1 hour per day). All experts had at least three years of experience in their respective fields. All participants were informed about the purpose of the study, and written consent was obtained to participate in the study. The module draft with all domains, activities/suggestions, and delivery methods of parent education was shared with the identified experts for their judgment. A google form was shared along with the module via email, and the participants were requested to review the module and respond to the google form having ten questions with a five-point rating scale related to the appropriateness of using the module in preschool children and an open-ended question where the participants could provide any suggestion or concerns related to the draft module.

### Parent feedback

The draft module was modified based on the expert’s inputs and suggestions and given to four parents of typically developing preschool children to receive their feedback. The parents were informed about the study purpose, and written consents were taken from them before they participated in the study. A Google (feedback) form, along with the revised module, was shared with the parents. The feedback form had four 5-point Likert scale questions, and an open-ended question. The participants were asked to check the module for satisfaction with content, duration, mode of delivery and acceptance.

## Results

The results of the study are presented under the following sections:

### Content development

A total of six parents participated in the in-depth interview for content development. The demographic data of all participants are provided in Table 1. A semi-structured interview was conducted on these participants using an interview guide with four open-ended questions. The parent’s responses were recorded, transcribed and coded to identify themes relevant to the module. Three themes and corresponding sub-themes were identified as presented in Table 2.

**Table 1.** Demographic details of parents (and their children) who participated in the in-depth interview.

| Participant | Age Range (years) | Gender | Child's age (years) | Child's Gender | Screen time (per day) |
| --- | --- | --- | --- | --- | --- |
| 1. | 36 - 40 | Male | 2.1 – 3.0 | Male | 4-5 hours |
| 2. | 36 - 40 | Female | 2.1 – 3.0 | Female | 3-4 hours |
| 3. | 36 - 40 | Male | 4.1 – 5.0 | Male | 3-4 hours |
| 4. | 31 - 35 | Female | 3.1 – 4.0 | Male | 1-2 hours |
| 5. | 31 - 35 | Female | 4.1 – 5.0 | Female | 2-3 hours |

**Table 2.**
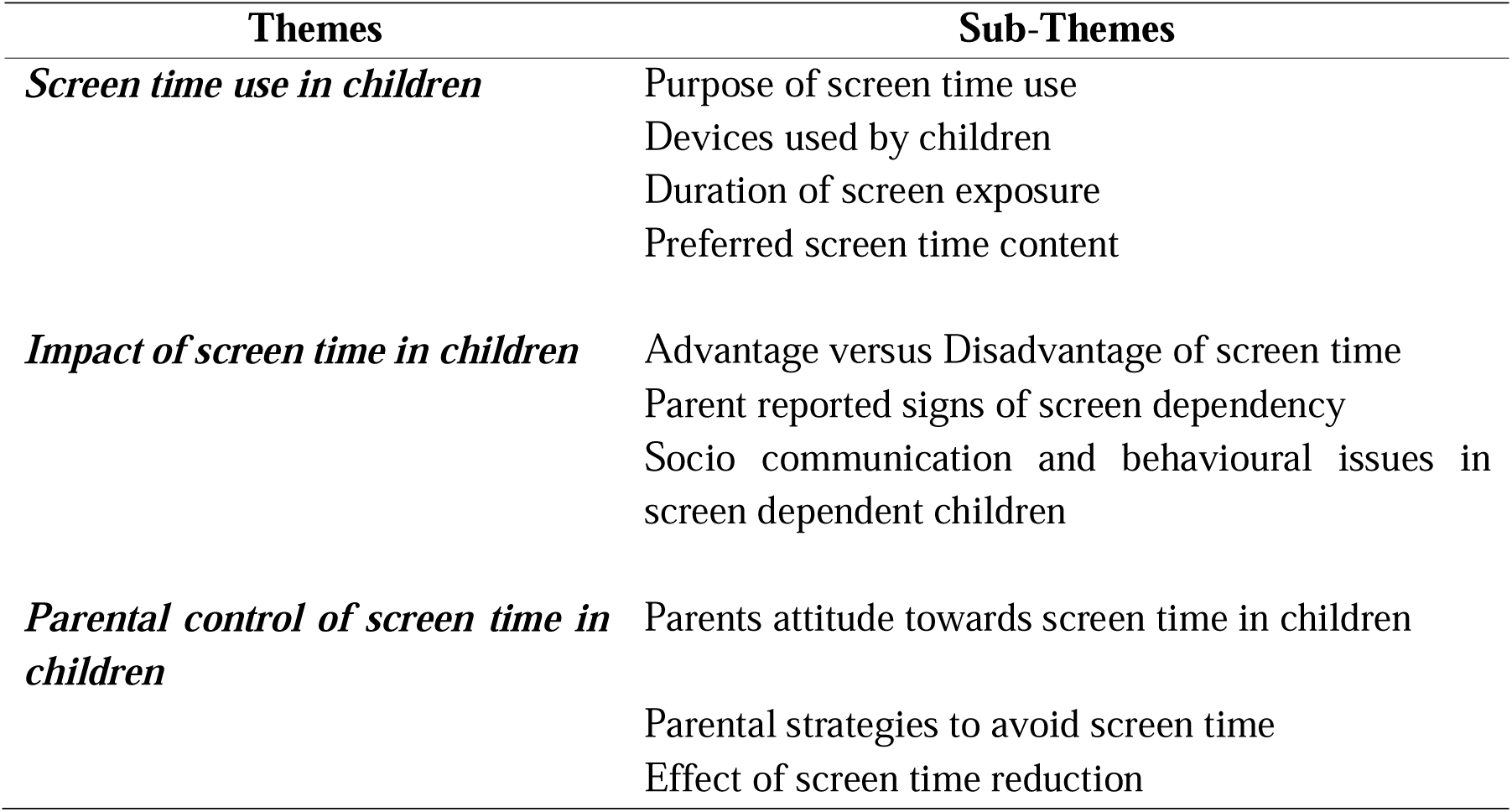
Themes and sub-themes derived from an in-depth interview conducted on the parents of children with excessive screen time.

| Themes | Sub-Themes |
| --- | --- |
| *Screen time use in children* | Purpose of screen time use<br>Devices used by children<br>Duration of screen exposure<br>Preferred screen time content |
| *Impact of screen time in children* | Advantage versus Disadvantage of screen time<br>Parent reported signs of screen dependency<br>Socio communication and behavioural issues in screen dependent children |
| *Parental control of screen time in children* | Parents attitude towards screen time in children<br>Parental strategies to avoid screen time<br>Effect of screen time reduction |

Based on the interview conducted on the parents, the following inferences were made: a) Most of the parents provide screen to their children during meals as well as to engage children while parents are unable to attend to them; b) Devices used by children during screen time were mostly mobile phones and television; c) Duration of screen exposure in the children averaged between 1.5 to 4 hours daily; d) Preferred screen content in children were cartoons and television advertisements.

Few parents favoured screen time in children, suggesting advantages such as – improvement in child’s vocabulary, supporting imitation skills, improvement in cognition and behaviour, and social communication skills in children. Parent’s feedback on the negative effects of excess screen time in children included speech delay, impact on the child’s brain development, effects on the child’s attention, concentration, and social skills. Parent-reported signs of screen dependency were behavioural issues (hitting, throwing objects, shouting, crying), irritability, temper tantrums when the screen was withdrawn, unable to shift the viewing until the program finished, and refusal to eat food. Socio-communication and behavioural issues reported in screen-dependent children were not responding to name calls, being short-tempered, crying and screaming, not showing interest in playing with peers, absence of social smile, poor eye contact, throwing objects to show anger, poor expressive language skills, and delayed language acquisition.

Most of the parents felt that screen time in children has a negative effect on their children. Parental strategies to avoid screen time included engaging them in outdoor activities, gardening, cycling, colouring and providing toys of the child’s interest. Few parents tried to reduce screen time in their children and reported improved behaviour, eye contact, attention, social smile, response to name call, and play behaviour.

### Item generation and Content validation

Based on the themes derived from the in-depth interview such as impact of screen time in children (social communication deficits), parental concerns regarding screen time in children, as well as literature search on available parent educational modules, a draft of parental educational module to reduce screen time and promote social communication skills in preschool children with excess screen time was prepared. The module draft was shared with healthcare professionals from various disciplines for content validation. Six experts reviewed the module (details of whom are provided in Table 3). These experts rated the contents of the draft module in terms of relevance, age appropriateness, feasibility, etc., using a 5-point Likert scale. The experts were also asked to provide comments/inputs related to the contents in the module.

**Table 3.** Demographic details of the experts who participated in content validation.

| Expert | Gender | Experience<br>(in years) | Work<br>setting | Education | Speciality |
| --- | --- | --- | --- | --- | --- |
| 1. | Female | 10 | Academic | Graduate | Speech and Language |
| 2. | Female | 5 | Hospital | Graduate | Paediatrics |
| 3. | Female | 7 | Clinical | Graduate | Occupational Therapy |
| 4. | Female | 6 | Clinical | Graduate | Psychology |
| 5. | Male | 26 | Hospital | Graduate | Psychiatry |
| 6. | Female | NA | IT | Undergraduate | Parent |

Experts rating was analysed to obtain the content validity index (CVI) in terms of S-CVI. The S-CVI value was 0.97, indicating the module is acceptable. For each of the ten questions, the item-level content validity index (I-CVI) is presented in Table 4. The overall agreement between the experts was estimated to be 80%, and the module is statistically acceptable.

**Table 4.** Results of content validity index calculation of the module.

| Statement(s) | Experts in<br>Agreement | I-CVI | Universal<br>Agreement<br>(UA) |
| --- | --- | --- | --- |
| The module is relevant to the current scenario of increased screen time in children. | 6 | 1 | 1 |
| Contents are in agreement with the current knowledge. | 6 | 1 | 1 |
| Contents are meeting the purpose of the intervention module. | 6 | 1 | 1 |
| The structure of the module is appropriate and convincing. | 6 | 1 | 1 |
| The duration of the programme is adequate. | 5 | 0.83 | 0 |
| The frequency of sessions is appropriate. | 6 | 1 | 1 |
| It is feasible to administer the programme to English-speaking Indian parents? | 6 | 1 | 1 |
| The module can be administered in online mode. | 6 | 1 | 1 |
| Listed general activities mentioned are appropriate for children between 2- 6 years of age. | 5 | 0.83 | 0 |
| Listed activities to improve social communication is appropriate for children between 2-6 years of age. | 6 | 1 | 1 |

Furthermore, experts provided several suggestions to improve the module for better effectiveness in the Indian context. They recommended including activities that are culturally relevant, adding more outdoor activities, and creating specific activities for children who seek sensory experiences. Additionally, incorporating pictures and visuals could help parents understand the content better. Some experts suggested increasing the duration and frequency of the sessions and offering additional online support to address any questions that might arise after the sessions. Other recommendations included making the module available in multiple Indian languages, delivering it in an online format, and incorporating online discussions, video demonstrations, and downloadable resources. Most of these suggestions were incorporated into the initial module, and a revised module was made. The final version of the parent educational module consisted of four sessions weekly once schedule of the parent educational contents using age-appropriate activities and plan to combat excess screen time and to support social communication skills in preschool children (Table 5). The four sessions addressed awareness of screen-time effects in children, behavioural issues due to screen time, social communication difficulties due to screen time and effective methods of screen use, along with follow-up discussions.

**Table 5.**
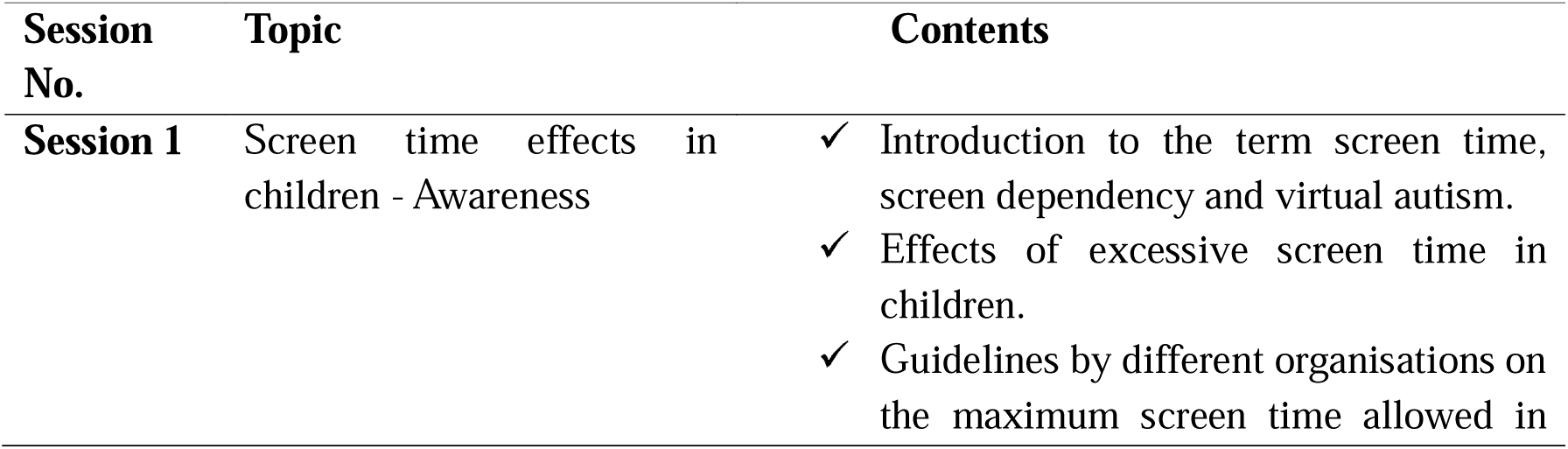
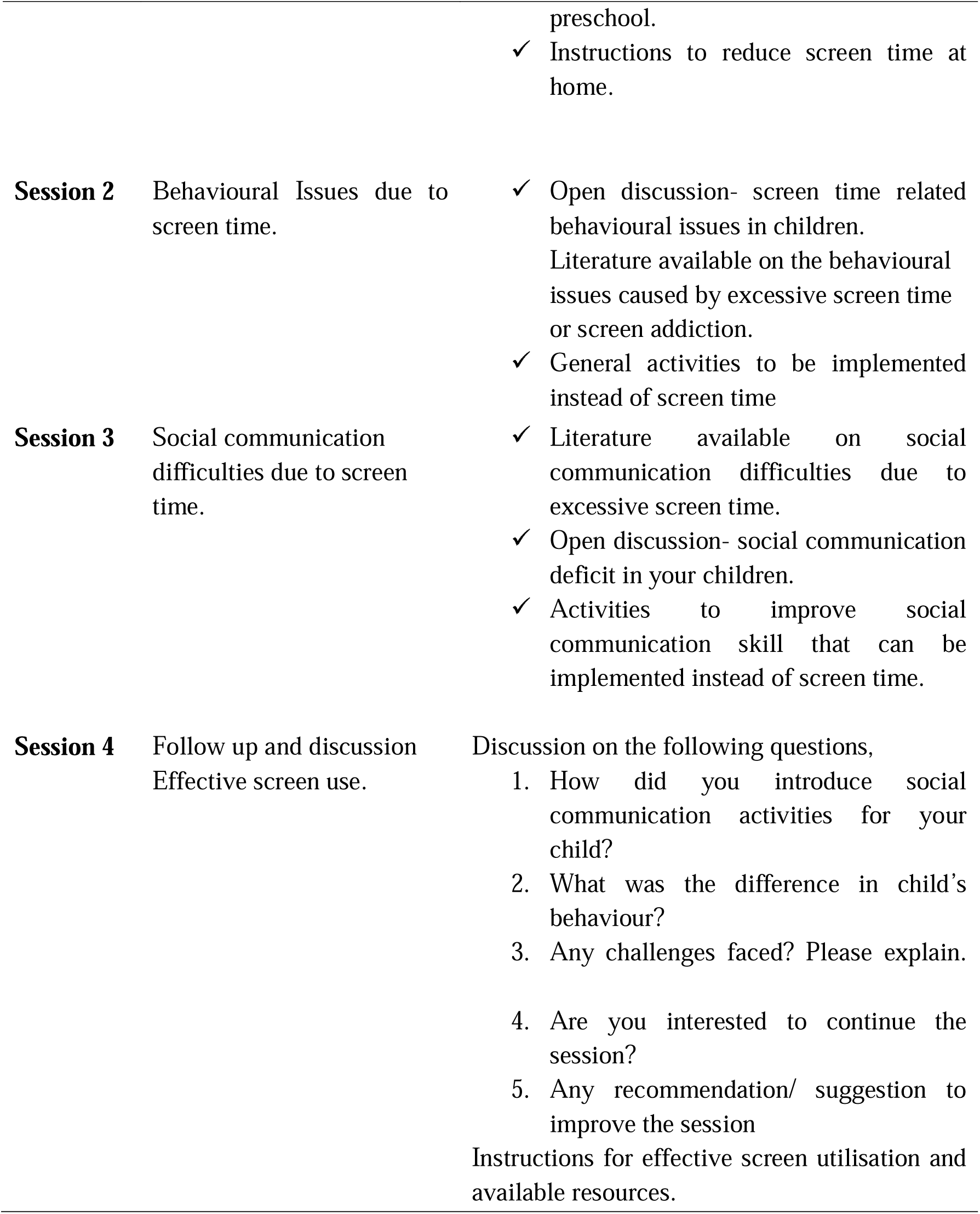
An overview of session-wise (weekly) contents for the parent educational module.

| Session No. | Topic | Contents |
| --- | --- | --- |
| **Session 1** | Screen time effects in children - Awareness | ✓ Introduction to the term screen time, screen dependency and virtual autism.<br>✓ Effects of excessive screen time in children.<br>✓ Guidelines by different organisations on the maximum screen time allowed in |
|  |  | preschool. |
|  |  | ✓ Instructions to reduce screen time at home. |
| **Session 2** | Behavioural Issues due to screen time. | ✓ Open discussion- screen time related behavioural issues in children.<br>Literature available on the behavioural issues caused by excessive screen time or screen addiction.<br>✓ General activities to be implemented instead of screen time |
| **Session 3** | Social communication difficulties due to screen time. | ✓ Literature available on social communication difficulties due to excessive screen time.<br>✓ Open discussion- social communication deficit in your children.<br>✓ Activities to improve social communication skill that can be implemented instead of screen time. |
| **Session 4** | Follow up and discussion<br>Effective screen use. | Discussion on the following questions,<br>1. How did you introduce social communication activities for your child?<br>2. What was the difference in child's behaviour?<br>3. Any challenges faced? Please explain.<br>4. Are you interested to continue the session?<br>5. Any recommendation/ suggestion to improve the session<br>Instructions for effective screen utilisation and available resources. |

### Parent Feedback

The revised parent educational module was then given to four parents to receive feedback on its relevance and utility. The feedback form consisted of 5 Likert scale questions and an open-ended question. The demographic details of the parents who provided feedback are presented in Table 6.

**Table 6.** Demographic details of parents who provided feedback on the educational module.

| Parent | Age (in years) | Gender | Education | Child's Age (in years) | Child's Gender |
| --- | --- | --- | --- | --- | --- |
| 1. | 26 - 30 | Female | Undergraduate | 4.1 – 5.0 | Female |
| 2. | 31 - 35 | Female | Undergraduate | 3.1 – 4.0 | Male |
| 3. | 36 - 40 | Male | Undergraduate | 2.1 – 3.0 | Male |
| 4. | 26 - 30 | Female | Undergraduate | 5.1 – 6.0 | Male |

The feedback revealed that around 75% of the parents were satisfied with the overall module, while one parent (25%) was not. Further, 75% of the parents felt that the module could help in reducing excess screen time in children. The module duration was perceived as satisfactory by 3 parents, while one parent gave a neutral response. Three of four parents agreed on the applicability of the activities in a home setting, although one parent disagreed. Regarding delivery of the therapy module, three of four parents preferred offline, while one parent preferred online mode. Further suggestions received by the parents were to provide materials or resources for the activities recommended and to include more interactive outdoor activities like going on a drive, engaging in extracurricular activities like musical school, martial arts, etc. These suggestions were further incorporated into the parent educational module.

## Discussion

Technology and digital media are developing faster than ever across the world. Technological advancements in various domains have drastically improved human quality of life in recent years and are expected to improve further. The need and use of technology have, in turn, increased the screen dependency in humans of all ages. A variety of screen devices are used by individuals for education, work, entertainment, communication, etc. Ever since COVID-19, the world has observed a significant rise in screen time, especially in children. This causes screen addiction, which has resulted in several health-related and behavioural issues in children. Excess screen time also causes developmental impairments (such as cognition and communication) and poor social skills. While there are guidelines available now on screen time limits for children of different ages, most of the children are exposed to screen time significantly higher than the recommended limits. More recent concept of virtual autism is thus proposed to refer to children with excessive screen time who present with behaviours and characteristics that mimic ASD. Children with such traits significantly improve their social and language skills when screen time is reduced with adequate guidance and stimulation. Thus, against the background of increased screen time in young children and a rise in related social communication difficulties, there is a need to develop parental educational programs addressing the need to reduce screen time and address the resulting social communication skills in such children. The present study, therefore, aimed to develop a parent educational module for preschool children (aged 2-5 years) who have excess screen time and experience social and communication issues.

An initial draft of the module was prepared, taking content from a focused literature search and conducting an in-depth interview with parents of children with increased screen time. The developed module was then shared with experts from various healthcare specialities (child psychiatrist, clinical psychologist, paediatrician, speech-language pathologist, occupational therapist), and a parent of preschool child, to receive their suggestions and ratings on the module’s contents. Based on the expert inputs, modifications were made to the module, and the updated module was used for parental feedback. Upon receiving the parental feedback, a few more changes were made in the module for finalization.

The results obtained from the semi-structured, in-depth interview on the parents of preschool children with excess screen time, in this study, revealed three main themes (viz., screen time use in children, impact of screen time in children, and parental control of screen time in children), and their corresponding sub-themes. Parental response to screen time issues reported in our study agreed with the available reports [42, 43]. Many parents use screens to keep their children occupied during meals or when they can’t pay direct attention to them. Children typically watch mobile phones and television, with cartoons and advertisements as the most popular content. Many parents expressed concerns about the negative impacts of screens, such as speech delays, hindered brain development, and difficulties with attention, concentration, and social skills [43]. Some parents reported behavioural issues, such as a lack of response to their names, irritability, poor eye contact, and delayed language skills in their children, which they believed was due to excessive screen time. To manage the screen time, parents often encouraged outdoor activities like gardening, cycling, and colouring, as well as provided toys that interest their children. Parents generally believed that screen time negatively affects their children, while a few of them reported improvements in their child’s behaviour when the screen exposure was reduced [44]. Some parents, rather reported potential benefits of screen use, including enhancements in vocabulary, imitation skills, cognition, and social communication [45]. The results of content validation of the developed module favoured acceptance of the module, showing a high degree of consensus among the raters and adequacy of the contents in the developed module. In addition, results of the parental feedback supported acceptance of the module for a majority (75%) of the participants. The parent educational module developed in this study, therefore, demonstrates adequate content as well as acceptance by the parents, and compares with similar intervention programs to combat excess screen time in preschool children [35, 37]. Boonmun et al. [37] developed a parent training program to reduce screen time in typically developing preschool children and documented its effectiveness in reducing screen time in Thailand. Kaur et al. [35] developed a parent training module for typical children with increased screen time in India. Subsequently, the authors [36] evaluated the intervention in parents and children where the child was within 2 to 5 years of age and found that it is effective in reducing screen time and increasing the duration of physical activity in children.

There are reports of a few parent training programs as being effective in reducing screen time in children with ASD as well. For instance, Khoo et al. [46] developed a parent training for children with ASD with increased screen time, in Malaysia. The training program was effective in reducing screen time and improving social skills. Heffler et al. [29] developed a parental module to reduce screen time in ASD children between 1.5 and 3.5 years of age. The module was effective in reducing screen time in children with autism. Further, the results also demonstrated reduced autistic behaviour as a secondary effect. Sadeghi et al. [34] conducted a parent training for parents of ASD children between 2-4 years of age and documented a reduction in screen time and stereotypic behaviour following training.

### Strengths and limitations

The present study developed a parental educational module using a rigorous method based on parental needs (gathered through in-depth interview) by combining expert opinion and parental feedback. Inputs from multiple disciplines were considered for the development of the module, making it developmentally appropriate. The inclusion of a parent in the expert panel is a step towards meeting the needs of the end user and making the module relevant. The home-based activities for the module were carefully selected by referring to the guidelines by the American Academy of Paediatrics (AAP), the World Health Organisation (WHO), and relevant literature. In addition to the available parent education programs to reduce screen time in children, this module includes activities and strategies to facilitate social communication skills in preschool children. The module could be administered online or offline and helps facilitate one-to-one interaction between the trainer and the parent. Additional support through online groups is expected to create a community of parents with similar concerns and provide help in sharing their concerns and strategies.

Screen time-related issues are a common concern to people of all ages. However, this module focuses on the screen time issues in preschool children only. The module in its current stage is not tested for its efficacy in reducing screen time and improving social communication skills in preschool children.

### Implications and future direction

The developed module holds the potential to be a promising educational program to reduce screen time in children and address its effects on their social communication behaviour. It also has the potential to improve awareness, knowledge and attitude of the parents towards screen time in preschool children. Future studies could validate the module by administering on parents of preschool children and determine its effect on reducing screen time in children and improving their social communication skills.

## Conclusion

The present study aimed to develop a parent educational module to meet the needs of screen-dependent, typically developing preschool children with social communication deficits. At first, the contents of the module were developed through a parental interview conducted to find the issues faced by parents due to screen time in children, in addition to a literature search. A panel of experts rated the contents of the module in terms of relevance, age appropriateness and feasibility of implementation, which showed high consensus among the raters and appropriate contents. The revised module’s expert suggestions further revealed acceptance by the parents. Activities specified in this module present a blend of age-appropriate physical, sensory, creative, imaginative, social play, and literacy skills, promoting fun and learning in the children. The module is expected to improve the parental awareness, knowledge and attitude about the challenges of screen time in children and thereby control the effects of screen time in this group.

## Data Availability

Data are available from the corresponding author upon request.

## Acknowledgement

The authors would like to express sincere gratitude to all the parents who participated in this study, as well as the experts who contributed their expertise for the content validation.

## Appendix

### Appendix-A Interview guide

Interview guide
| Sl No | Interview question |
| --- | --- |
| **Q1** | Please provide details of your child's gadget use on an average daily basis. (types, duration, purpose and frequency of gadget use) |
| **Q2** | What is your opinion about children's use/experience of gadgets, who are under five years of age? |
| **Q3** | Do you think that as a parent you need to monitor your child's use of electronic gadgets? (If so, why, how, what is your child's response to restrictions in gadget use, how do you overcome that, what are the issues or challenges faced in restricting gadget use in your child) |
| **Q4** | Does your child experience any social or communication difficulties? Please elaborate on the nature, type and severity of these issues in comparison to children of his/her age. |

